# Prevalence and determinants of persistent symptoms after infection with SARS-CoV-2: Protocol for an observational cohort study (LongCOVID-study)

**DOI:** 10.1101/2022.01.10.22269009

**Authors:** Elizabeth N. Mutubuki, Tessa van der Maaden, Ka Yin Leung, Albert Wong, Anna D. Tulen, Siméon de Bruijn, Lotte Haverman, Hans Knoop, Eelco Franz, Albert Jan van Hoek, Cees C. van den Wijngaard

**Author notes:** **Corresponding author:** Elizabeth N Mutubuki.

## Abstract

**Background:** A substantial proportion of individuals infected with severe acute respiratory syndrome coronavirus-2 (SARS-CoV-2) report persisting symptoms weeks and months following acute infection. Estimates on prevalence vary due to differences in study designs, populations, heterogeneity of symptoms and the way symptoms are measured. Common symptoms include fatigue, cognitive impairment and dyspnea. However, knowledge regarding the nature and risk factors for developing persisting symptoms is still limited. Hence in this study we aim to determine the prevalence, severity, risk factors and impact on quality of life of persisting symptoms in the first year following acute SARS-CoV-2 infection.

**Methods:** The LongCOVID-study is both a prospective and retrospective cohort study with a one year follow up. Participants aged 5 years and above with self-reported positive or negative tests for SARS-CoV-2 will be included in the study. The primary outcome is the prevalence and severity of persistent symptoms in participants that tested positive for SARS-CoV-2 compared to controls. Symptom severity will be assessed for fatigue using the Checklist Individual Strength (CIS subscale fatigue severity), pain (Rand-36/SF-36 subscale bodily pain), dyspnea (Medical Research Council (mMRC)) and cognitive impairment using the Cognitive Failure Questionnaire (CFQ). Secondary outcomes include loss of health-related quality of life (HRQoL) and risk factors for persisting symptoms following infection with SARS-CoV-2.

**Discussion:** A better understanding regarding the nature of persisting symptoms following SARS-CoV-2 infection will enable better diagnosis, management and will consequently minimize negative consequences on quality of life.

## Background

During the first months of the pandemic, epidemiological research focused primarily on the spread of Severe Acute Respiratory Syndrome Coronavirus-2 (SARS-CoV-2) and on treatment of those with severe or fatal illness (1). The effects of SARS-CoV-2 infection vary from asymptomatic infection, through to critical and chronic disease (2). Although most individuals infected with SARS-CoV-2 fully recover, there is a growing body of evidence that suggests that a substantial number of individuals remain with long-term complications or persisting symptoms (3-5).

COVID-19 varies in clinical presentation, disease severity, recovery time as well as completeness of recovery (6). A delay in recovery whereby individuals fail to return to their normal daily routines and still report lasting effects of the infection long after the expected period of recovery has been termed ‘‘LongCOVID’’ (7), ‘‘long-haulers” (8) and “post COVID-19 condition” (9). The term post COVID-19 condition will be used in the rest of this article. Post COVID-19 condition is reported to occur in individuals that have a history of probable or confirmed SARS-CoV-2 infection, usually 3 months from the onset of COVID-19 and symptoms with a duration of at least 2 months that cannot be explained by alternative diagnosis (9). Fatigue, shortness of breath, cognitive dysfunction are some of the common symptoms (9). Symptoms may persist from initial infection, be a new onset following initial recovery from an acute COVID-19 episode or may also fluctuate or relapse over time (9).

Over 210 million confirmed cases of COVID-19 have been reported, and of those, an estimated 10-20% are reported to experience such persisting symptoms for weeks and months following acute SARS-CoV2 infection (9). However, higher incidence rates of persisting symptoms have been reported, for example through self-surveys of patient from long COVID peer support groups (10) as well as in hospitalized patients (11). Variation in the reported incidence and prevalence rates of post COVID-19 condition can be attributed to the complexity of the syndrome, differences in population groups, heterogeneity in clinical presentation of symptoms, little knowledge regarding the natural history and clinical course (12) and in the way symptoms are measured. Common persistent symptoms are shortness of breath, fatigue, dyspnea and headaches (13, 5). Some of the initial acute symptoms such as cough, fever, and chills become less prevalent as the illness progresses, whereas cognitive dysfunction and palpitations become more prevalent later in the illness (5).

A good overview of the nature of persisting symptoms following an acute infection with SARS-CoV-2, can enable better diagnosis, management and may reduce negative consequences on HRQoL (12). Hence in this study we aim to determine the prevalence and severity of persisting symptoms in the first year of infection, in individuals infected by SARS-CoV-2 compared to individuals that were not infected. In addition risk factors for developing post COVID-19 condition and its impact on health will be analyzed.

## Methods/Design

### Study aim and design

The LongCOVID-study is an observational cohort study consisting of prospective and retrospective data with one year of follow up. The study aims to determine the prevalence, severity, health impact and risk factors associated with persistent symptoms following a SARS-CoV-2 infection, in cases compared to population controls and test-negative controls. The study is carried out by the Dutch National Institute for Public Health and the Environment (RIVM), Bilthoven, the Netherlands. The Utrecht Medical Ethics Committee (METC) declared in February 2021 that the Medical Research Involving Human Subjects Act (WMO) does not apply to this study (METC protocol number 21-124/C).

### Study population

Both the prospective and retrospective cohorts include children (ages 5-17) and adults (18 years and above).

#### Prospective cohort study

Participants with a positive SARS-CoV-2 infection test result on an antigen or polymerase chain reaction (PCR) test for acute infection, are included in the study as cases, if they complete the baseline questionnaire within 7 days of testing positive. Participants that test negative to SARS-CoV-2 infection and complete their baseline questionnaire within 7 days of testing negative, are included in the study as test-negative controls. A second group of controls, population controls, consists of randomly selected participants from the Basic Registration of Persons (BRP) without a positive test for SARS-CoV-2 infection or known history of probable infections.

#### Retrospective cohort study

Participants presenting with self-reported persisting symptoms associated with SARS-CoV-2 infection with or without having had a positive test result were included in the retrospective cohort study as post COVID-19 condition cases.

### Recruitment

Figure 1 shows the flow diagram of participant recruitment in the LongCOVID-study. Participants are recruited through the following three ways;

**Figure 1:**
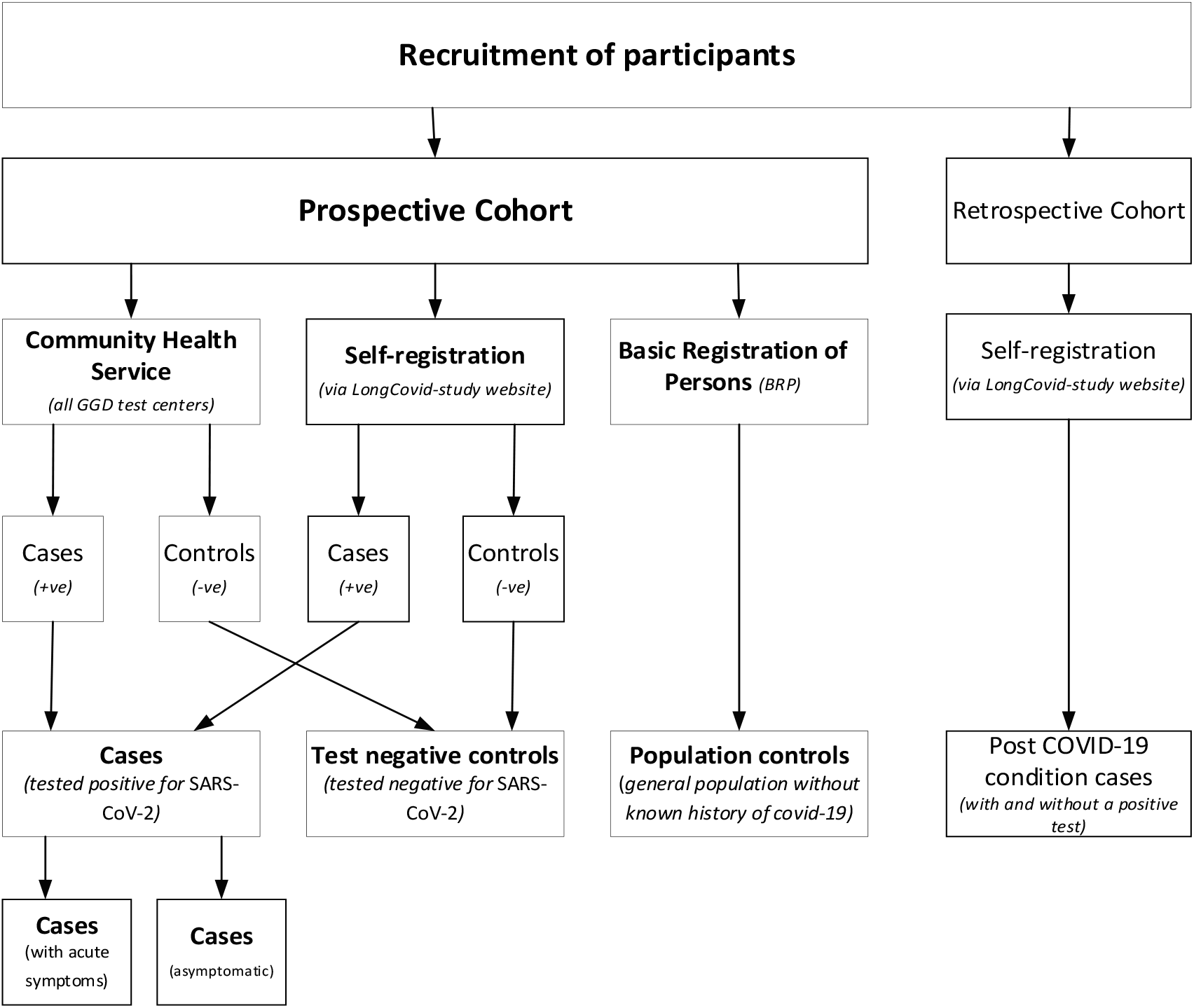
Recruitment of participants in the LongCOVID-study

#### Via the Community Health Service

Individuals testing positive and negative to COVID-19 at one of the community health services (GGDs) are invited to participate in the LongCOVID-study. Registration to participate is via the LongCOVID-study website.

#### Basic Registration of Persons (BRP)

Population controls are randomly selected from the basic registration of persons and invited by letter to participate in the study.

#### Self-registered participants

Individuals interested in participating in the LongCovid-study can also self-register through the study website (longcovid.rivm.nl). Test-negative controls, cases and post COVID-19 condition cases can be included in the study this way.

### Measurements (adults)

Table 1 shows different measurement moments were data is collected in form of questionnaires. At baseline, data on demographical characteristics such as gender, education level and employment are collected. Data on comorbidities is reported at baseline and at 12 months. Information regarding testing for SARS-CoV-2, COVID-19 related complaints and vaccination data is collected at baseline and at 3, 6, 9 and 12 months.

**Table 1:**
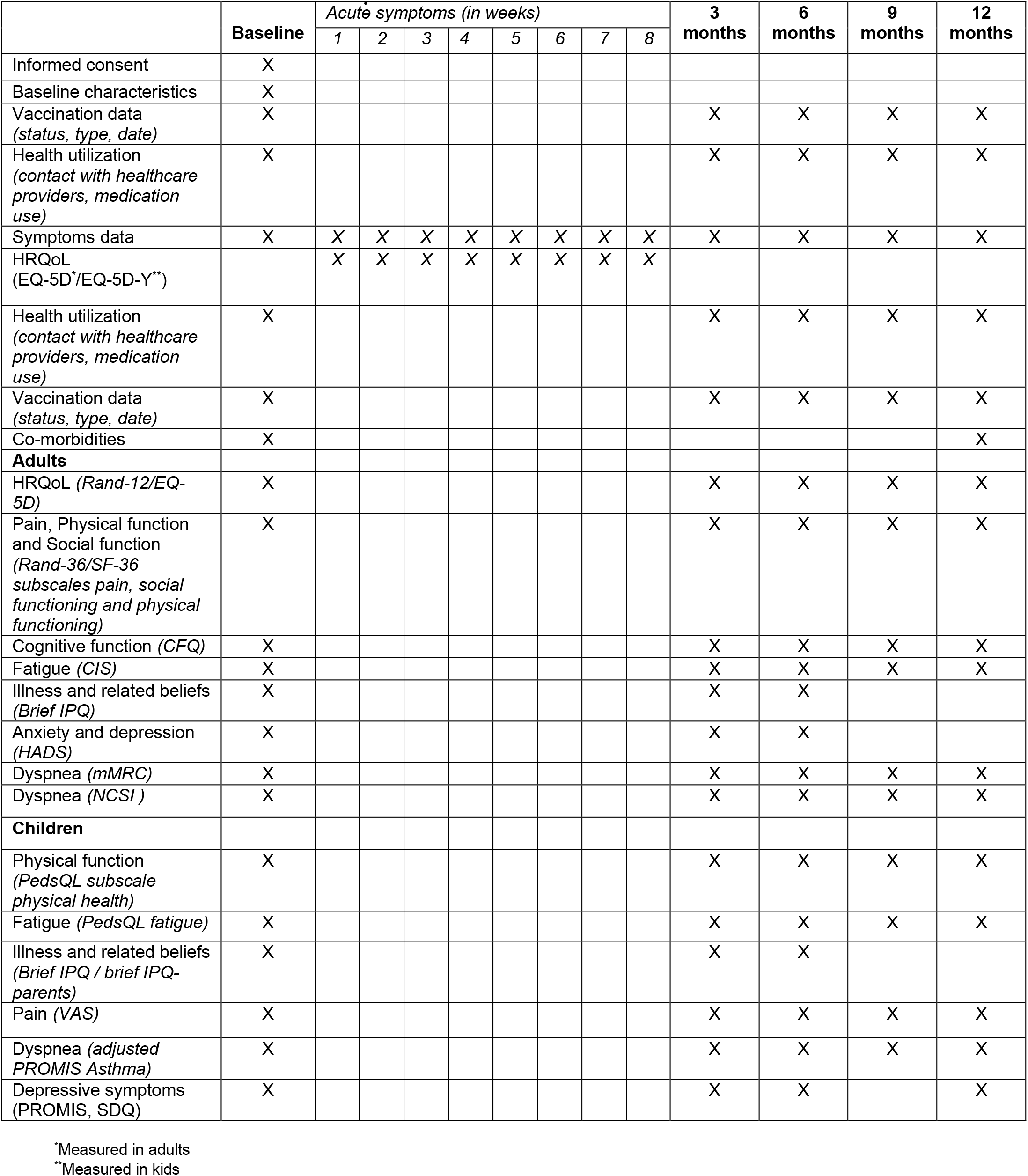
Measurement moments

|  | Baseline | Acute symptoms (in weeks) |  |  |  |  |  |  |  | 3 months | 6 months | 9 months | 12 months |
| --- | --- | --- | --- | --- | --- | --- | --- | --- | --- | --- | --- | --- | --- |
|  |  | 1 | 2 | 3 | 4 | 5 | 6 | 7 | 8 |  |  |  |  |
| Informed consent | X |  |  |  |  |  |  |  |  |  |  |  |  |
| Baseline characteristics | X |  |  |  |  |  |  |  |  |  |  |  |  |
| Vaccination data<br>(status, type, date) | X |  |  |  |  |  |  |  |  | X | X | X | X |
| Health utilization<br>(contact with healthcare providers, medication use) | X |  |  |  |  |  |  |  |  | X | X | X | X |
| Symptoms data | X | X | X | X | X | X | X | X | X | X | X | X | X |
| HRQoL<br>(EQ-5D*/EQ-5D-Y**) |  | X | X | X | X | X | X | X | X |  |  |  |  |
| Health utilization<br>(contact with healthcare providers, medication use) | X |  |  |  |  |  |  |  |  | X | X | X | X |
| Vaccination data<br>(status, type, date) | X |  |  |  |  |  |  |  |  | X | X | X | X |
| Co-morbidities | X |  |  |  |  |  |  |  |  |  |  |  | X |
| **Adults** |  |  |  |  |  |  |  |  |  |  |  |  |  |
| HRQoL (Rand-12/EQ-5D) | X |  |  |  |  |  |  |  |  | X | X | X | X |
| Pain, Physical function and Social function<br>(Rand-36/SF-36 subscales pain, social functioning and physical functioning) | X |  |  |  |  |  |  |  |  | X | X | X | X |
| Cognitive function (CFQ) | X |  |  |  |  |  |  |  |  | X | X | X | X |
| Fatigue (CIS) | X |  |  |  |  |  |  |  |  | X | X | X | X |
| Illness and related beliefs<br>(Brief IPQ) | X |  |  |  |  |  |  |  |  | X | X |  |  |
| Anxiety and depression<br>(HADS) | X |  |  |  |  |  |  |  |  | X | X |  |  |
| Dyspnea (mMRC) | X |  |  |  |  |  |  |  |  | X | X | X | X |
| Dyspnea (NCSI ) | X |  |  |  |  |  |  |  |  | X | X | X | X |
| **Children** |  |  |  |  |  |  |  |  |  |  |  |  |  |
| Physical function<br>(PedsQL subscale physical health) | X |  |  |  |  |  |  |  |  | X | X | X | X |
| Fatigue (PedsQL fatigue) | X |  |  |  |  |  |  |  |  | X | X | X | X |
| Illness and related beliefs<br>(Brief IPQ / brief IPQ-parents) | X |  |  |  |  |  |  |  |  | X | X |  |  |
| Pain (VAS) | X |  |  |  |  |  |  |  |  | X | X | X | X |
| Dyspnea (adjusted PROMIS Asthma) | X |  |  |  |  |  |  |  |  | X | X | X | X |
| Depressive symptoms<br>(PROMIS, SDQ) | X |  |  |  |  |  |  |  |  | X | X |  | X |

\*Measured in adults
\*\*Measured in kids

#### Health related quality of life (EQ-5D-5L and Rand-12/SF-12)

HRQoL is assessed using the Rand-12/SF-12 and EQ-5D-5L. Additional weekly measurement using the EQ-5D-5L were carried out in individuals presenting with acute symptoms in the first 8 weeks following a positive COVID-19 test. The EQ-5D-5L questionnaire consists of five dimensions of health (mobility, self-care, usual activities, pain/discomfort, and anxiety/ depression), with five levels of response and a visual analogue scale (EQ VAS). The EQ-5D-5L scores will be converted into utility scores using the Dutch tariff (14), ranging from 0 (death) to 1 (optimal health).

The Rand-12/SF-12, a shortened version of the Rand-36/SF-36 HRQoL questionnaire consists of 12 questions from the following 8 domains; physical functioning, physical role, emotional role limitations, social functioning, physical pain, general mental health, vitality and general health perception. The 8 domains can be summarized into a physical and mental health domain (15). Health scores will be converted into utility scores using the SF-6D (Short-Form Six-Dimension). Quality adjusted life years will be calculated by multiplying the utility scores by the time a patient spends in a given health state.

#### Fatigue (Checklist Individual Strength [CIS])

Fatigue severity is assessed using the subscale fatigue severity of the Checklist Individual Strength (CIS). The CIS subscale fatigue is a 8-item fatigue questionnaire (16). Each item is scored on a 7-point Likert scale. Scores range from 8 to 56, and scores of 35 and higher indicate severe fatigue (17).

#### Cognitive function (Cognitive Failure Questionnaire [CFQ])

Cognitive function is assessed using the Cognitive Failure Questionnaire (CFQ). The CFQ ranges from 0 to a 100 with higher scores indicating more cognitive impairment (18). A score of 44 or higher indicated clinically significant complaints on cognitive function.

#### Pain (bodily pain subscale of the Rand-36/SF-36 Health Status Inventory [Rand-36])

The bodily pain subscale of the Rand-36 Health Status Inventory (Rand-36) is used to assess pain severity. The Rand-36 scores range from 0 to 100, higher scores indicate better health status. Significant impairment due to pain is reflected by a score of 55 or lower, based on Dutch norm scores (19). The subscales physical and social functioning were also used.

#### Dyspnea (Medical Research Council (dyspnea) [mMRC])

Dyspnea is assessed using the modified Medical Research Council (dyspnea) (mMRC). The mMRC scale ranges from grade 0 to 4. Grade 0-breathless with strenuous exercise; Grade 1-short of breath when hurrying on level ground or walking up a slight hill; Grade-2 walks slower on level ground because of breathlessness or stops for a breath when walking at own pace; Grade 3-stops for breath after walking about 100 yards or after a few minutes on level ground and Grade 4-am too breathless to leave the house or I am breathless when dressing (20). A score of 1 or higher reflecting significant impairment due to dyspnea (21).

#### Illness and related beliefs (The Brief Illness Perception Questionnaire [Brief IPQ])

The Brief Illness Perception Questionnaire (Brief IPQ /IPQ-K) is a nine-item scale to assess the cognitive and emotional representations of illness including consequences, timeline, personal control, treatment control, identity, coherence, concern, emotional response and causes (22). Item scores increases, represent linear increases in the dimension measured. The Brief IPQ is reported to have good test-retest reliability (22).

#### Anxiety (Hospital Anxiety and Depression Scale [HADS])

HADS (Hospital Anxiety and Depression Scale) is a 14 item self-report questionnaire designed to measure anxious and depressive states in patients with two subscales (23). The sum score per subscale ranges from 0 to 21. Scores between 0-7 indicate no anxiety or depression, 8-10 mild cases, 11-15 moderate cases and 16 or above severe cases (Snaith 1994).

#### Dyspnea (The Nijmegen Clinical Screening Instrument [NCS])

The Nijmegen Clinical Screening Instrument (NCSI) measures health status and has the following domains, physiological functioning, symptoms, functional impairment, and quality of life as main domains (24), and 8 subdomains (25). The 8 subdomains include subjective symptoms, dyspnea emotions, fatigue, behavioral impairment, subjective impairment, general quality of life (general QoL), health related quality of life (HRQoL) and satisfaction with relations (25). Each subdomain is expressed as a single score on its own scale, with higher NCSI scores indicate more problems (24). In the study the subdomain dyspnea will be used.

#### Absenteeism (iMTA Productivity Cost Questionnaire [iPCQ])

Participants were asked to report the number of days that they had been absent from work due to illness. Absenteeism will be measured using the iPCQ.

#### Unpaid Productivity losses and informal care

Unpaid productivity losses from work, studies, voluntary work as well as informal care will be valued using the Dutch shadow price of 14,57 euros per hour (26).

### Measurements (children)

Below are age-specific scales that were used in children (aged 5-17 years), that replaced some of the above described scales for adults (Table 1).

#### Physical function (Pediatric Quality of Life Inventory [PedsQL])

The PedsQL is a HRQoL measure consisting of 4 subscales (physical functioning, emotional functioning, social functioning and school functioning) which can be computed to two summary scores (psychosocial and physical health summary scores).Dutch norms are available which allow comparison with the general population (27). A parent proxy of the PedsQL will be used for children aged 5-7 years.

#### Fatigue (Pediatric Quality of Life Inventory Fatigue Scale [PedsQL fatigue])

Fatigue severity in children will be assessed with the Pediatric Quality of Life Inventory Fatigue Scale (PedsQL fatigue). This 18-item PedsQL fatigue scale comprises the general fatigue scale (6 items), sleep/rest fatigue scale (6 items), and cognitive fatigue scale (6 items) and is a reliable and valid instrument to measure fatigue in children (28). Dutch norm scores are available (29). A parent proxy will be used for children aged 5-7 years.

#### Illness and related beliefs (The Brief Illness Perception Questionnaire [Brief IPQ])

The Brief Illness Perception Questionnaire (Brief IPQ /IPQ-K-parents) will be completed by a proxy (22), and by the child if they are aged 10 or older.

#### Pain Visual analogue scale (VAS)

Pain severity will be assessed using VAS (30). Scores range from 0 (no pain) to a 100 (worst imaginable pain). A parent proxy will be used for children aged 5-7 years.

#### Health related quality of life (EQ-5D-Y)

Weekly measurement moments for up to 8 weeks in children presenting with acute symptoms follow a positive COVID-19 test, will be carried out. The EQ-5D-Y-Proxy1 will be used for children aged 5-7 years, and the EQ-5D-Y will be used for children aged 8-17 years to measure HRQoL. The EQ-5D-Y-Proxy1 and EQ-5D-Y questionnaires consist of five dimensions (mobility, self-care, usual activities, pain/discomfort, and anxiety/ depression), with three levels of response and a visual analogue scale (EQ VAS)(31).

#### Dyspnea (Patient-Reported Outcomes Measurement Information System [PROMIS])

Dyspnea will be assessed in children aged 5-7 years using an adjusted Patient-Reported Outcomes Measurement Information System (PROMIS) Asthma impact Short form Proxy and in children 8-17 using an adjusted PROMIS Asthma impact Short form(32).

#### Cognitive function and behaviour (PROMIS and SDQ)

Loneliness will be assessed in children aged 5-7 years using the PROMIS short form proxy depressive symptoms and in children 8-17 using the PROMIS short form depressive symptoms, for which norm scores are available which allow comparison with the general population (32, 33). In addition the strengths and difficulties questionnaire (SDQ) will be used as well to assess the level of depressive symptoms, with a proxy for parents in the 5-11 years of age (34).

### Measurements (acute cohort)

Data on HRQoL and acute symptoms will be collected weekly in the first 8 weeks following infection. Data collection will stop when the symptoms stop or end at 8 weeks following infection.

### Outcome measures

#### Primary outcome

1. The first primary outcome measure is the prevalence and severity of persistent symptoms in patients that tested positive for COVID-19 infection compared to both test-negative controls and population controls. Severity of symptoms will be assessed for fatigue, pain, dyspnea, and cognitive impairment using standardized questionnaires, with population-based norm cut-off scores for clinically significant severity.

#### Secondary outcomes include

1. Factors that predict post COVID-19 condition following an acute SARS-CoV-2 infection at different follow up moments.
2. Healthcare utilization in the first year following infection with SARS-CoV-2 in cases compared to controls (test-negative controls and population controls) will be assessed.
3. Health related quality of life in cases will be compared to that of controls (test-negative controls and population controls) in the first year following infection.

Additionally a comparison will also be made between post COVID-19 condition individuals and individuals that test positive for COVID-19 but do not develop post COVID-19 condition.

### Statistical analysis

Baseline characteristics of the participants in all groups will be presented using descriptive statistics mean (standard deviation), median (range), or proportion to assess if there is a balance in the groups regarding distribution of prognostic factors such as age, gender, co-morbidity and education.

### Prospective study

#### Primary outcome analysis

##### 1. Prevalence and severity of persistent symptoms in COVID-19 patients

Descriptive epidemiological statistical methods will be used to analyze prevalence of persistent symptoms at 3, 6, 9 and 12 months in cases compared to both control groups (test-negative controls and population controls). Persisting symptoms are defined as symptoms in cases with a duration of at least 2 months. Such symptoms markedly elevated in cases compared to controls (test-negative controls and/or population controls) during follow up are likely to be associated with COVID-19, and cases with these symptoms are in this study defined as cases with possible post COVID-19 condition (yes/no). Severity scores of fatigue, dyspnea, cognitive functioning, and pain will be calculated. Scores of individuals with confirmed COVID-19 will be compared to those of controls, per follow-up moment (baseline, 3, 6, 9 and 12 months follow-up).

#### Secondary outcome analysis

##### 1. Predictors of post-COVID 19 condition

A prediction model will be built to identify predictors of post COVID-19 condition. The outcome will be having possible post COVID-19 condition as defined above. To determine the prediction model that best suits our data, the prediction model will be constructed using super learning (35). The prediction model will be evaluated using the ROC-AUC metric (36).

##### 2. Predictors of healthcare utilization in post COVID-19 condition

A second prediction model will be performed to identify predictors of healthcare utilization in post COVID-19 condition. Healthcare utilization is defined as contact (visit to the general practitioner, telephone call, hospitalization, emergency healthcare services, other medical health professionals/services) with a health provider regarding symptoms attributed by the patient to COVID-19 or post COVID-19 condition (yes/no). The prediction model will be performed as mentioned above.

##### 3. Quality-adjusted life-years

HRQoL will be assessed using EQ-5D-5L and Rand-12/SF-6D. Quality-adjusted life-years (QALYs), which takes into account both the impact of length and the quality-of-life will be calculated and be compared between cases and controls.

### Retrospective study

Descriptive epidemiological statistical methods will be used to analyze the prevalence of persistent symptoms at baseline in cases compared to both control groups (test-negative controls and population controls). Moreover, prevalence of co-morbidities will be quantified in cases and control groups. Additionally, an assessment into healthcare utilization for cases will be performed according to the aforementioned definition.

### Acute data following infection

Descriptive epidemiological statistical methods will be used to describe the prevalence and the type of symptoms present following acute infection as well as health related quality of life.

#### Missing data

The fraction of missing questionnaires at each time points and per period during the study (e.g., per 3 months) in all patients with confirmed Covid-19 will be tabulated. Scenarios of dealing with missing data include a complete case analysis, multiple imputation, and linear interpolation combined with carry forward.

## Discussion

The LongCOVID-study aims to determine the prevalence and severity of persistent symptoms following acute SARS-CoV2 infection in cases compared to controls, as well as to investigate the risk factors of developing persistent symptoms. Previous studies have explored prevalence of long-term symptoms and risk factors in various populations, i.e., in previously hospitalized patients (37), patients with diabetes type 1 and 2 (38, 39), in home-isolated patients with milder symptoms and in the young (40).

Blomberg reported that 61% of all the patients had persisting symptoms at 6 months (40). This included patients with a mild to moderate illness following initial illness as well as young patients (16-30 years). Persisting symptoms included loss of taste and or smell, fatigue, dyspnea, impaired concentration and memory problems. In a hospitalized population (37), fatigue, muscle weakness, sleep difficulties and anxiety or depression are the most prevalent symptoms at 6 months. Due to severe illness during hospital stay and impaired pulmonary function, the hospitalized population is a target group for long-term recovery (37, 41). Our study includes both adults and children from the age of 5 with mostly mild to moderate acute symptoms and a much smaller group of patients that were hospitalized in the acute phase of the infection. We expect a possible bias against the number of hospitalized patients due to the design of the study, which requires questionnaires to be completed no more than seven days following a positive test for COVID-19.

Strengths of the current study include the prospective design, allowing for detailed analysis of the prevalence and risk factors of persistent symptoms of SARS CoV-2 infection. In addition this study is one of a few studies (42) that allows for comparison of COVID-19 cases to control groups that have similar experiences, such as lock down measures. This is important because such factors can influence complaints. The availability of the population control group in this study allows us to control for background prevalence of symptoms. In addition the use of test-negative controls allows for assessment of the impact of COVID-19 compared to other respiratory infections. The use of validated questionnaires with validated cut-off scores for severity is another strength of this study. Repeated assessment of symptoms every three months during one year of follow-up will enable assessment of the time course of symptoms, and detection of disabling symptoms at every 3 months interval.

Furthermore, the impact of symptoms on general functioning will be assessed. A limitation of this study is that severity scores of only four of symptoms associated with COVID-19 will be calculated to get more insight into clinical significance. This is because only four standardized questionnaires for symptom severity were included in the study. Hence the severity of other possible symptoms related to COVID-19 will not be taken into account.

In conclusion, the LongCOVID-study is expected to provide additional insights into the prevalence and severity of persistent symptoms after SARS CoV-2 infection to the international body of literature. In the Netherlands this is the first large scale study on persisting symptoms following SARS CoV2 infection.

## Data Availability

This article is a protocol for an observational cohort study, hence data sharing is not applicable as no datasets were generated or analyzed during the current study.

## Declarations

### Ethics approval and consent to participate

All participants give consent to participate in the study. Written consent is obtained online prior to being able to complete the study questionnaires, which are also completed online. The Utrecht Medical Ethics Committee (METC) declared in February 2021 that the Medical Research Involving Human Subjects Act (WMO) does not apply to this study (METC protocol number 21-124/C).

### Consent for publication

Not applicable

### Availability of data and materials

Data sharing is not applicable to this article as no datasets were generated or analyzed during the current study.

### Authors’ contributions

ENM wrote the manuscript. KYL contributed to the methods. CCW, TM, AJH, ADT, KYL, AW contributed to the design of the study, LH and HK advised on the design of the questionnaires, TM, AJH, ADT, KYL, ENM, EF, SB, CCW contributed to implementation of the data collection, all authors reviewed and contributed to drafts of the manuscript. All authors read, contributed to refinement of the study protocol and approved the manuscript.

## Acknowledgements

Not applicable

## List of abbreviations

SARS-CoV-2: Severe Acute Respiratory Syndrome Coronavirus-2
CIS: Checklist Individual Strength
mMRC: Medical Research Council
HRQoL: Health-Related Quality of Life
RIVM: Dutch National Institute for Public Health and the Environment
METC: Medical Ethics Review Committee
WMO: Medical Research Involving Human Subjects Act
PCR: Polymerase Chain Reaction
BRP: Basic Registration of Persons
GGD: Municipal Health Services
CFQ: Cognitive Failure Questionnaire
Brief IPQ: The Brief Illness Perception Questionnaire
HADS: Hospital Anxiety and Depression Scale
NCSI: Nijmegen Clinical Screening Instrument
iPCQ: iMTA Productivity Cost Questionnaire
PedsQL: Pediatric Quality of Life Inventory
PedsQL fatigue: Pediatric Quality of Life Inventory Fatigue Scale
VAS: Visual Analogue Scale
PROMIS: Patient-Reported Outcomes Measurement Information System
QALYs: Quality Adjusted Life Years
SDQ: Strengths and Difficulties Questionnaire

